# Vaccine-related misinformation and attitude toward COVID-19 vaccination in Japan

**DOI:** 10.1101/2024.07.17.24310600

**Authors:** Yuki Furuse, Takahiro Tabuchi

## Abstract

We have struggled with vaccine hesitancy for vaccination rollout during the COVID-19 pandemic. Although the spread of misinformation seems to play a role in vaccine hesitancy, the extent to which it affects attitudes toward COVID-19 vaccination is unknown. Here, we investigated the prevalence of beliefs about misinformation regarding COVID-19 vaccines in vaccinated and unvaccinated populations in Japan and analyzed associated risk factors. An online survey of 31,000 participants in 2021 found that 8.1% of vaccine-accepted individuals believed vaccine-related misinformation, whereas 36.6% of those who refused vaccination believed misinformation. Most factors associated with beliefs about misinformation and vaccine hesitancy overlapped, including young age, unmarried status, low income, particular information sources, and history of COVID-19 infection. Interestingly, some factors, such as age and sources of information, had different effects on vaccine acceptance between individuals who did not believe misinformation and those who did. Advanced age was associated with vaccine acceptance among non-misinformation believers. In contrast, misinformation believers in their 10s and 20s were more willing to be vaccinated than older adults. The effects of television and Internet information were stronger in individuals who believed misinformation on their attitude toward vaccination than non-misinformation believers. This study highlights the importance of understanding the relationship between beliefs about misinformation and vaccine hesitancy for ongoing and future pandemics.

## Background

The COVID-19 pandemic has caused substantial morbidity and mortality worldwide. To reduce the death toll, novel vaccines have been developed and administered (1). Vaccination for COVID-19 in Japan started in February 2021. During the time, “one million shots per day” was aimed by the Japanese government to increase vaccination coverage as high and as rapidly as possible (2).

However, we have faced vaccine hesitancy at that time. Many people are especially concerned about the effectiveness and safety of mRNA vaccines, a new biomedical technology that had not been practically applied before the pandemic (3). Furthermore, misinformation regarding COVID-19 and its vaccines has spread worldwide, dubbed an infodemic (4,5). Those include unfounded allegations like “governments hide the causal association between vaccination and autism” or “the COVID-19 pandemic was caused by a man-made virus to control the global population by vaccination.” Such misinformation has been reported to be associated with vaccine hesitancy (6).

Still, some people must have received vaccination even if they believed vaccine-related misinformation. How often that happened and what determined the behavior are largely unknown. In this study, we investigated the prevalence of people who believed misinformation regarding COVID-19 vaccines by vaccination status, factors associated with misinformation beliefs, and how they have affected attitudes toward COVID-19 vaccination in Japan.

## Methods

### Survey for vaccine hesitancy and beliefs about misinformation

Data on vaccine hesitancy and beliefs about vaccine-related misinformation were collected in part of the Japan COVID-19 and Society Internet Survey (JACSIS) study (7). Briefly, 31,000 participants aged 15 years or older were recruited and asked about their demographic, economic, and health-related information through an online questionnaire between September and October 2021. Participants were asked whether they agreed with each of the seven sentences regarding vaccine-related misinformation (Table 1).

**Table 1.** Seven vaccine-related misinformation sentences asked in JACSIS.

|  |
| --- |
| Misinformation regarding COVID-19 vaccines |
| 1. The reported data about vaccine safety are fabricated. |
| 2. Vaccines for infants and children are harmful, but the fact is concealed. |
| 3. Pharmaceutical companies hide safety concerns in vaccines. |
| 4. Citizens are deceived about the effectiveness of vaccines. |
| 5. The reported data about vaccine effectiveness are fabricated. |
| 6. Citizens are deceived about the safety of vaccines. |
| 7. Governments hide the causal association between vaccination and autism. |

Responses from each participant were weighted using inverse probability weighting for participation in the JACSIS study to be adjusted to the general population in Japan, referring to the national data of the Comprehensive Survey of Living Conditions in 2019. Propensity scores were calculated by adjusting for residential address (prefecture), education level, marital status, and self-rated health consciousness (8,9).

### Factors for vaccine hesitancy and misinformation beliefs

Factors associated with vaccine hesitancy were tested with binomial regression analysis using the Stats package in R. Similarly, factors associated with the number of beliefs about vaccine-related misinformation were tested by ordered logistic regression analysis using the MASS package in R. Statistical adjustments were conducted for age groups where indicated. Statistical significance was set at P < 0.05.

### Ethical consideration

The protocol for the JACSIS study was reviewed and approved by the Institutional Review Board of the Osaka International Cancer Institute (No. 20084). The survey results were anonymized, and all study participants agreed that their response data would be used for analysis and publication.

## Results

Of the 31,000 participants in the JACSIS study in 2021, 28,175 (90.9 %) provided valid responses (7). Among them, 83.2% (23,435) were already vaccinated for COVID-19, 3.4% (968) were waiting for vaccination, 6.2% (1,740) were undecided about whether to get vaccinated, 6.4% (1,800) decided not to get vaccinated, and 0.8% (232) responded that they could not get vaccinated for medical reasons, such as allergies (Table 2).

**Table 2.** Summary of JACSIS study about vaccine hesitancy and beliefs about vaccine-related misinformation.

|  |  |  |  | Vaccine acceptance |  |  |  |  | Number of believed vaccine-related misinformation items |  |  |  |  |  |  |  |
| --- | --- | --- | --- | --- | --- | --- | --- | --- | --- | --- | --- | --- | --- | --- | --- | --- |
| Age group | All participants | Valid responders | Percent tage of valid responders | Already vaccinated | Want to be vaccinated (waiting) | Wondering | Refuse to get vaccinated | Unable to get vaccinated because of medical reasons | 0 | 1 | 2 | 3 | 4 | 5 | 6 | 7 |
| 10 | 645 | 573 | 88.8% | 434 (75.7%) | 37 (6.5%) | 44 (7.7%) | 52 (9.1%) | 6 (1.0%) | 471 (82.2%) | 53 (9.2%) | 24 (4.2%) | 11 (1.9%) | 6 (1.0%) | 2 (0.3%) | 3 (0.5%) | 3 (0.5%) |
| 20 | 4,107 | 3,626 | 88.3% | 2,767 (76.3%) | 223 (6.2%) | 307 (8.5%) | 305 (8.4%) | 24 (0.7%) | 3,055 (84.3%) | 286 (7.9%) | 127 (3.5%) | 45 (1.2%) | 31 (0.9%) | 28 (0.8%) | 17 (0.5%) | 37 (1.0%) |
| 30 | 4,652 | 4,149 | 89.2% | 3,077 (74.2%) | 270 (6.5%) | 398 (9.6%) | 371 (8.9%) | 33 (0.8%) | 3,589 (86.5%) | 231 (5.6%) | 99 (2.4%) | 61 (1.5%) | 24 (0.6%) | 32 (0.8%) | 39 (0.9%) | 74 (1.8%) |
| 40 | 6,078 | 5,454 | 89.7% | 4,330 (79.4%) | 240 (4.4%) | 430 (7.9%) | 388 (7.1%) | 66 (1.2%) | 4,798 (88.0%) | 246 (4.5%) | 115 (2.1%) | 55 (1.0%) | 56 (1.0%) | 46 (0.8%) | 50 (0.9%) | 88 (1.6%) |
| 50 | 5,289 | 4,785 | 90.5% | 3,999 (83.6%) | 147 (3.1%) | 291 (6.1%) | 311 (6.5%) | 37 (0.8%) | 4,274 (89.3%) | 207 (4.3%) | 73 (1.5%) | 39 (0.8%) | 28 (0.6%) | 33 (0.7%) | 68 (1.4%) | 63 (1.3%) |
| 60 | 5,267 | 4,882 | 92.7% | 4441 (91.0%) | 35 (0.7%) | 160 (3.3%) | 209 (4.3%) | 37 (0.8%) | 4,489 (92.0%) | 177 (3.6%) | 49 (1.0%) | 36 (0.7%) | 19 (0.4%) | 26 (0.5%) | 28 (0.6%) | 58 (1.2%) |
| 70 | 4,962 | 4,706 | 94.8% | 4,387 (93.2%) | 16 (0.3%) | 110 (2.3%) | 164 (3.5%) | 29 (0.6%) | 4,365 (92.8%) | 162 (3.4%) | 46 (1.0%) | 27 (0.6%) | 31 (0.7%) | 23 (0.5%) | 22 (0.5%) | 30 (0.6%) |
| Total | 31,000 | 28,175 | 90.9% | 23,435 (83.2%) | 968 (3.4%) | 1,740 (6.2%) | 1,800 (6.4%) | 232 (0.8%) | 25,041 (88.9%) | 1,362 (4.8%) | 533 (1.9%) | 274 (1.0%) | 195 (0.7%) | 190 (0.7%) | 227 (0.8%) | 353 (1.3%) |

No vaccine-related misinformation was believed in 88.9% (25,041) of the participants. Among the already vaccinated people, 8.1% believed at least one of the seven misinformation items (Table 1, Figure 1A and Supplementary Figure 1). In contrast, 36.6% of those who refused vaccination believed misinformation. To put it the other way around, 88.1% of people who did not believe misinformation were willing to be vaccinated, while 63.6% of misinformation believers accepted vaccination (Figure 1B). Vaccine acceptance rates declined as the number of believed misinformation items increased.

**Figure 1.**
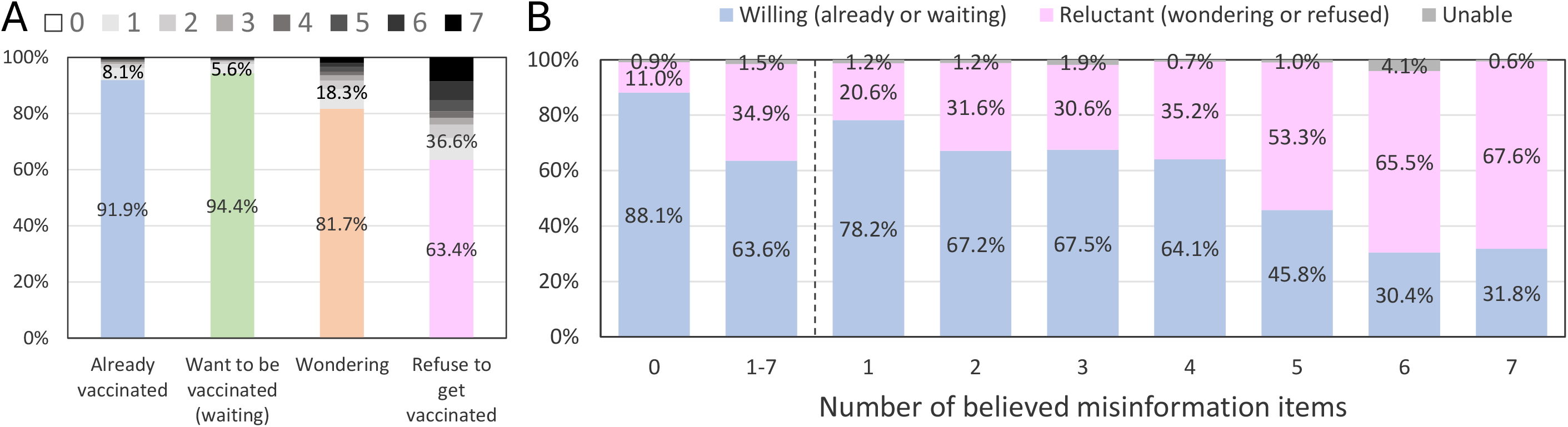
Prevalence of vaccine hesitancy and misinformation beliefs A) The prevalence of beliefs about vaccine-related misinformation was stratified by attitudes toward vaccination. B) The proportions of vaccine acceptance and hesitancy were shown by the number of believed misinformation items. “1–7” indicates people who believed at least one vaccine-related misinformation item. The survey results were adjusted for age, residence, educational level, marital status, and self-rated health consciousness to fit the general population in Japan. The results for each age group are shown in Supplementary Figure 1.

Younger age; living by oneself; unmarried; lower education level; not having physical comorbidity; having a mental illness; history of COVID-19 infection; not knowing friends who had contracted COVID-19 before; not getting information about COVID-19 and its vaccination from friends, medical doctors, governments, newspapers, or television; getting information from the Internet including social media; and lower income were significantly associated with vaccine hesitancy (Table 3). In addition, the number of believed vaccine-related misinformation items was strongly associated with vaccine hesitancy, with odds ratios ranging from 2.1 (believed one misinformation item) to 17.5 (believed seven misinformation items). Hence, many factors associated with vaccine hesitancy overlapped with those related to beliefs about misinformation. Younger age; living by oneself; unmarried; having a mental illness; history of COVID-19 infection; having family members and friends who had contracted COVID-19 before; not getting information from governments, newspapers, or television; getting information from the Internet; and lower income were significantly associated with an increase in the number of believed misinformation items (Table 4).

**Table 3.** Risk factors for vaccine hesitancy.

| Category | Variable | Vaccination |  | Unadjusted |  | Age-adjusted |  |
| --- | --- | --- | --- | --- | --- | --- | --- |
|  |  | Willing<br>(already or waiting) | Reluctant<br>(wondering or<br>refused) | Odds ratio | P-value | Odds ratio | P-value |
| Sex | Male | 12042 (86.8%) | 1744 (12.6%) | Reference |  | Reference |  |
|  | Female | 12361 (86.4%) | 1796 (12.6%) | 1.00 (0.93–1.08) | 0.9283 | 1.02 (0.95–1.1) | 0.5007 |
| Age | 15–19 | 471 (82.2%) | 96 (16.8%) | 1.14 (0.9–1.43) | 0.272 |  |  |
|  | 20–29 | 2990 (82.5%) | 612 (16.9%) | **1.14 (1.02–1.28)** | **0.0216** |  |  |
|  | 30–39 | 3347 (80.7%) | 769 (18.5%) | **1.28 (1.15–1.43)** | **<0.0001** |  |  |
|  | 40–49 | 4570 (83.8%) | 818 (15.0%) | Reference |  |  |  |
|  | 50–59 | 4146 (86.6%) | 602 (12.6%) | **0.81 (0.72–0.91)** | **<0.0001** |  |  |
|  | 60–69 | 4476 (91.7%) | 369 (7.6%) | **0.46 (0.40–0.52)** | **<0.0001** |  |  |
|  | 70– | 4403 (93.6%) | 274 (5.8%) | 0.35 (0.30–0.40) | 0.3695 |  |  |
| Living | By oneself | 4818 (81.5%) | 1027 (17.4%) | 1.66 (1.53–1.80) | <0.0001 | **1.59 (1.46–1.72)** | **<0.0001** |
| Marital status | Married | 15247 (89.9%) | 1604 (9.5%) | 0.50 (0.46–0.53) | <0.0001 | **0.57 (0.52–0.61)** | **<0.0001** |
| Education level | Junior high school | 276 (74.4%) | 86 (23.2%) | 2.07 (1.60–2.65) | <0.0001 | **2.23 (1.71–2.87)** | **<0.0001** |
|  | High school | 6224 (86.1%) | 938 (13.0%) | Reference |  | Reference |  |
|  | University/college | 14865 (87.7%) | 1957 (11.5%) | 0.87 (0.80–0.95) | 0.0015 | **0.74 (0.68–0.81)** | **<0.0001** |
|  | Graduate school | 1127 (89.9%) | 124 (9.9%) | 0.73 (0.60–0.89) | 0.0018 | **0.57 (0.46–0.69)** | **<0.0001** |
| Comorbidity | Physical illness | 14760 (88.4%) | 1760 (10.5%) | 0.65 (0.60–0.69) | <0.0001 | **0.78 (0.72–0.84)** | **<0.0001** |
|  | Mental illness | 1645 (81.4%) | 331 (16.4%) | 1.43 (1.26–1.61) | <0.0001 | **1.29 (1.14–1.46)** | **0.0001** |
| COVID-19 infection history | Self | 399 (77.9%) | 91 (17.8%) | 1.59 (1.25–1.99) | 0.0001 | **1.36 (1.02–1.80)** | **0.0318** |
|  | Family member | 424 (79.5%) | 84 (15.8%) | 1.37 (1.08–1.73) | 0.0084 | 1.09 (0.81–1.45) | 0.5503 |
|  | Friend | 2063 (86.4%) | 284 (11.9%) | 0.94 (0.83–1.07) | 0.3874 | **0.73 (0.63–0.83)** | **<0.0001** |
| Information source | Family member | 12638 (89.4%) | 1376 (9.7%) | 0.59 (0.55–0.64) | <0.0001 | 1.04 (0.95–1.13) | 0.443 |
|  | Friend | 13355 (89.8%) | 1406 (9.4%) | 0.55 (0.51–0.59) | <0.0001 | **0.70 (0.63–0.76)** | **<0.0001** |
|  | Medical doctor | 7799 (92.7%) | 529 (6.3%) | 0.37 (0.34–0.41) | <0.0001 | **0.60 (0.54–0.66)** | **<0.0001** |
|  | Expert | 10083 (90.5%) | 968 (8.7%) | 0.53 (0.49–0.58) | <0.0001 | 0.97 (0.89–1.07) | 0.5594 |
|  | Government | 11575 (91.1%) | 1019 (8.0%) | 0.45 (0.41–0.48) | <0.0001 | **0.66 (0.61–0.72)** | **<0.0001** |
|  | Internet including social media | 17411 (87.2%) | 2375 (11.9%) | 0.82 (0.76–0.88) | <0.0001 | **1.48 (1.35–1.62)** | **<0.0001** |
|  | Newspaper | 11218 (92.0%) | 883 (7.2%) | 0.39 (0.36–0.42) | <0.0001 | **0.69 (0.63–0.76)** | **<0.0001** |
|  | Television | 20429 (89.4%) | 2238 (9.8%) | 0.33 (0.31–0.36) | <0.0001 | **0.48 (0.43–0.52)** | **<0.0001** |
| Income | 0–3 million JPY | 3977 (81.7%) | 811 (16.7%) | 1.58 (1.43–1.74) | <0.0001 | **1.82 (1.64–2.02)** | **<0.0001** |
|  | 3–6 million JPY | 7593 (87.9%) | 982 (11.4%) | Reference |  | Reference |  |
|  | 6–9 million JPY | 4362 (89.3%) | 494 (10.1%) | 0.88 (0.78–0.98) | 0.0229 | **0.73 (0.65–0.82)** | **<0.0001** |
|  | 9–12 million JPY | 2078 (90.8%) | 198 (8.7%) | 0.74 (0.63–0.86) | 0.0002 | **0.62 (0.52–0.73)** | **<0.0001** |
|  | 12 million– JPY | 1300 (89.7%) | 138 (9.5%) | 0.82 (0.68–0.99) | 0.0391 | **0.72 (0.59–0.87)** | **0.0007** |
| Believed misinformation items | 0 | 22404 (89.5%) | 2464 (9.8%) | Reference |  | Reference |  |
|  | 1 | 1068 (78.4%) | 272 (20.0%) | 2.32 (2.01–2.66) | <0.0001 | **2.12 (1.84–2.44)** | **<0.0001** |
|  | 2 | 358 (67.2%) | 165 (31.0%) | 4.19 (3.46–5.05) | <0.0001 | **3.65 (3.01–4.42)** | **<0.0001** |
|  | 3 | 171 (62.4%) | 94 (34.3%) | 5.00 (3.86–6.43) | <0.0001 | **4.56 (3.51–5.90)** | **<0.0001** |
|  | 4 | 114 (58.5%) | 77 (39.5%) | 6.14 (4.57–8.21) | <0.0001 | **5.98 (4.43–8.04)** | **<0.0001** |
|  | 5 | 90 (47.4%) | 97 (51.1%) | 9.8 (7.33–13.11) | <0.0001 | **9.59 (7.13–12.91)** | **<0.0001** |
|  | 6 | 79 (34.8%) | 141 (62.1%) | 16.23 (12.32–21.53) | <0.0001 | **16.33 (12.33–21.78)** | **<0.0001** |
|  | 7 | 119 (33.7%) | 230 (65.2%) | 17.57 (14.06–22.07) | <0.0001 | **17.46 (13.91–22.03)** | **<0.0001** |
| Total |  | 24403 (86.6%) | 3540 (12.6%) |  |  |  |  |

Significance after age adjustment is in bold.

**Table 4.**
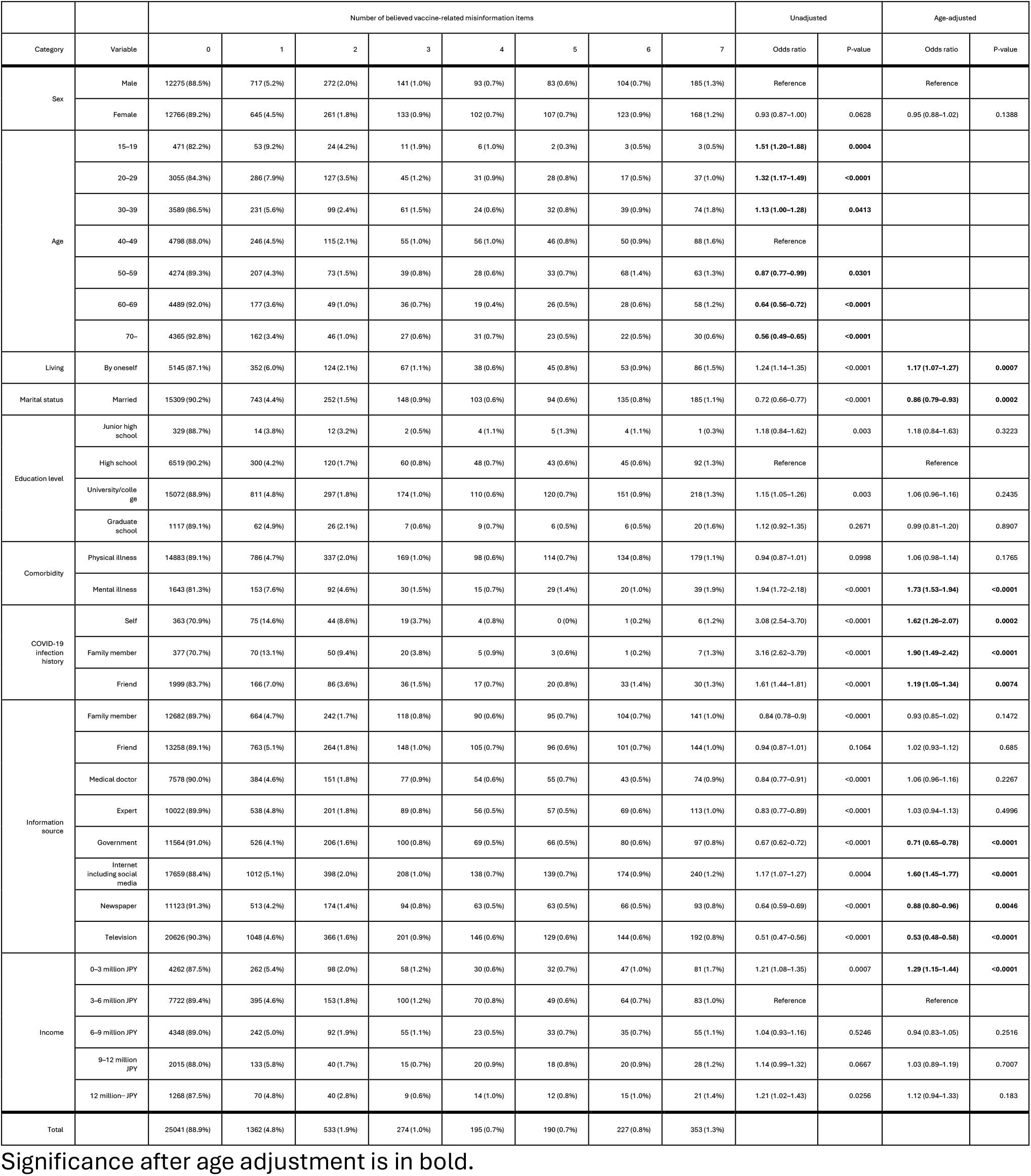
Risk factors for misinformation beliefs.

We noticed that many people who believed vaccine-related misinformation still accepted vaccination (Figure 1B) and wondered whether any factors affected their attitudes toward vaccination differently between individuals who did not believe misinformation and those who did. Figure 2 shows the odds ratios for vaccine hesitancy in the two stratified groups. Although the direction of the associations was the same for most factors between the two groups, two findings were notable. First, age had the opposite effect. Whereas older age showed a strong association with vaccine acceptance among non-misinformation believers, there was a positive association between younger age and vaccine acceptance among misinformation believers. Second, the effects of some information sources were more evident among individuals who believed misinformation. The association between an information source from television and vaccine acceptance was stronger among misinformation believers than non-misinformation believers. An information source from the Internet was found to be more closely linked to vaccine hesitancy in misinformation believers than in non-misinformation believers.

**Figure 2.**
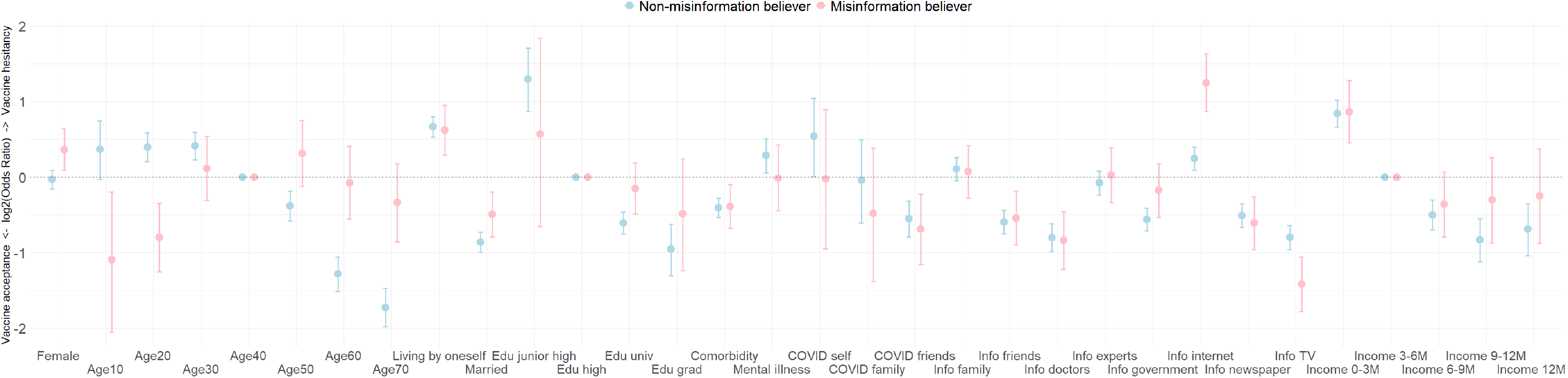
Factors associated with vaccine hesitancy in non-misinformation and misinformation believers The age-adjusted odds ratios for vaccine hesitancy in non-misinformation believers (zero believed misinformation items) and misinformation believers (2 or more believed misinformation items) are shown. Vertical lines indicate 95% confidence intervals.

## Discussion

In this study, we reported the prevalence of people who believed vaccine-related misinformation in vaccinated and unvaccinated populations in Japan through an online survey that included over 30,000 participants. We also identified the risk factors associated with misinformation beliefs and vaccine hesitancy.

The spread of misinformation and vaccine hesitancy are major hurdles in the fight against COVID-19. As with previous studies, we found that many factors, such as the absence of cohabiters, low income, particular information sources, and history of COVID-19 infection, are associated with misinformation beliefs and vaccination reluctance (10–12). An interesting observation of the present study is that the effect of misinformation on vaccine hesitancy is weak in the younger generation (Figure 2 and Supplementary Figure 1B), although COVID-19 vaccination coverage is generally high for older adults (1). While information from the Internet helps misinformation-believers have a more obstinate attitude toward vaccine hesitancy, information from television showed a positive effect on vaccine acceptance, even for individuals who believed vaccine-related misinformation. Further studies on these mechanisms will allow us to develop effective strategies to increase vaccination coverage in the midst of an infodemic.

Our study has several limitations. The generalizability of the results is limited, as we conducted the survey only for residents of Japan at a certain time. As the participants were recruited through an online platform, we cannot rule out the existence of selection bias. Although we statistically adjusted our data to the general population in the country, this could have been insufficient. In addition, the self-report of vaccination status in the survey was not verified by official records.

COVID-19 vaccination coverage in Japan is very high, reaching >80% of eligible people by the end of 2021 (1). We found that 88.9% of the study participants did not believe vaccine-related misinformation. In addition, among those who believed misinformation, 63.6% accepted vaccination. However, there is a growing trend of vaccine hesitancy today. For example, the coverage of regular vaccinations, such as one for measles, has recently decreased in Japan and other countries (13,14). As the spread of misinformation repeatedly hinders countermeasures for past infectious disease outbreaks (15), this tide should not be disregarded for the next pandemic. Our study underscores the importance of understanding risk factors for misinformation beliefs in relation to vaccine hesitancy.

## Data availability

All data produced in the present study are available upon reasonable request to the authors.

## Author contributions

TT was a major contributor to the JACSIS survey. YF analyzed and interpreted the data and wrote the first draft. All authors read and approved the final manuscript.

## Acknowledgments

This work was supported by the Strategic Center of Biomedical Advanced Vaccine Research and Development for Preparedness and Response (JP243fa627001 and JP243fa627004) from the Japan Agency for Medical Research and Development; by the Grant-in-Aid for Scientific Research (JP16KK0059, JP18H03107, JP19K10446, JP19KK0204, JP21H04856, and JP23K09693) from the Japan Society for the Promotion of Science; by the Health Labour Sciences Research Grants (19FA1012) from the Ministry of Health, Labour and Welfare; by the Innovative Research Program on Suicide Countermeasures (R3-2-2) from the Japan Suicide Countermeasures Promotion Center; by the Grants from Chiba Foundation for Health Promotion & Disease Prevention (grant number not available); and by the Nagasaki University State of the Art Research program from Nagasaki University (grant number not available). The funders had no role in the study design, data collection and analysis, decision to publish, or preparation of the manuscript.

