## Supplementary materials for "Vaccine-related misinformation and attitude toward COVID-19 vaccination in Japan"

### Supplementary Figures

A

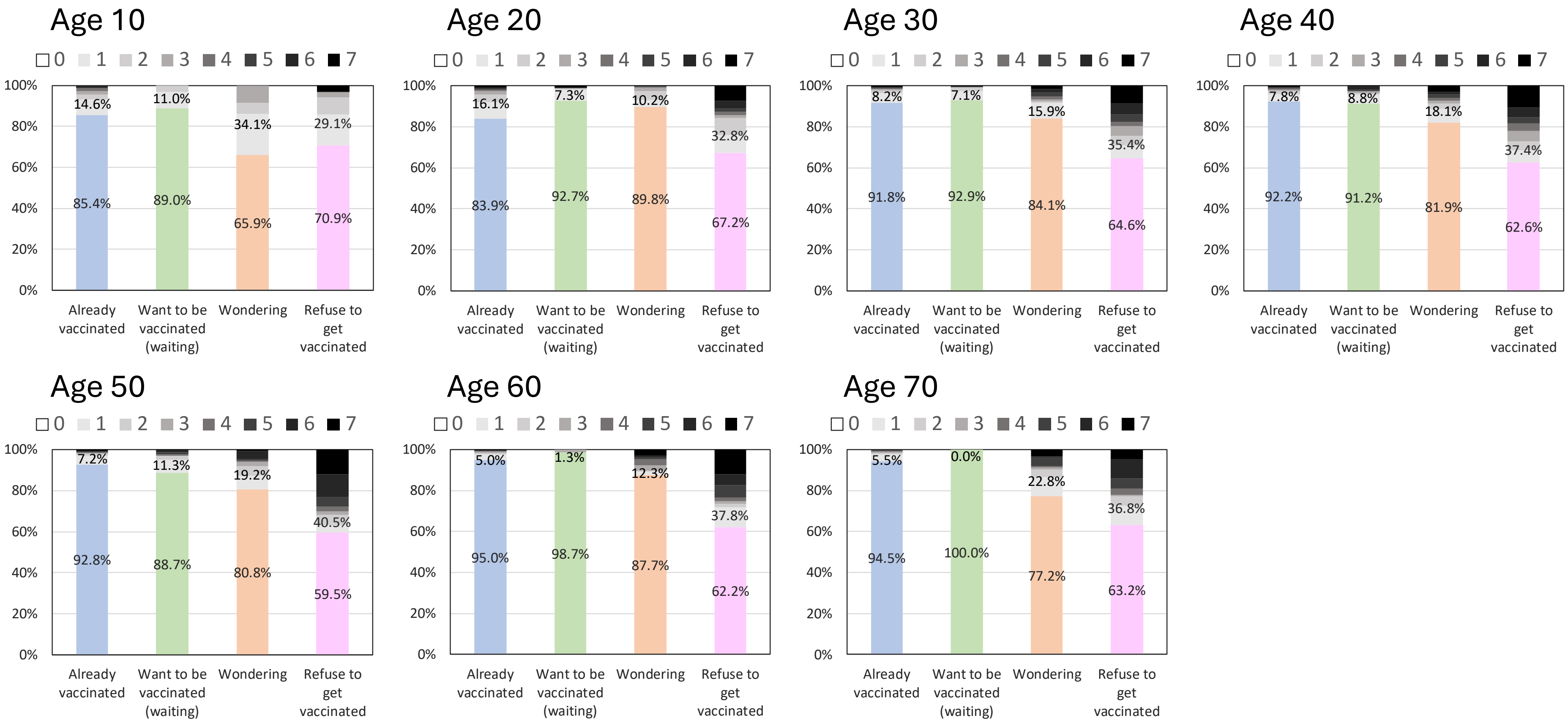

B

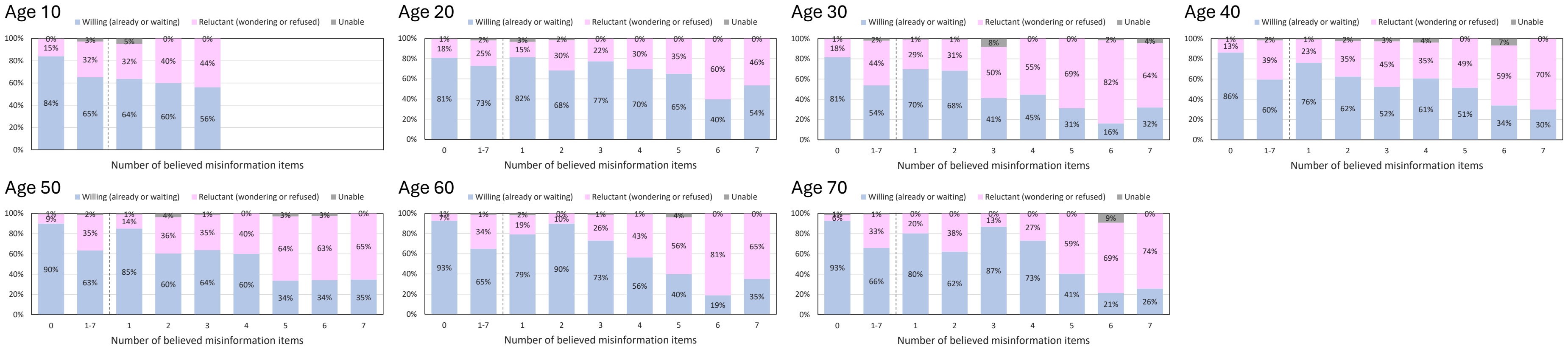


Supplementary Figure 1 Prevalence of vaccine hesitancy and beliefs about vaccine-related misinformation in each age group

A) Prevalence of misinformation beliefs stratified by attitude toward vaccination. B) Proportion of vaccine acceptance and hesitancy by the number of believed misinformation items. “1–7” indicates people who believed at least one vaccine-related misinformation item. For age group 10, graphs are not shown for those who believed four or more misinformation items due to a small number of <10.
